# Comparison of the therapeutic efficacy of antibiotic-loaded polymeric tissue scaffold and bone cement in the regeneration of infected bone tissue

**DOI:** 10.1101/2023.08.23.23294525

**Authors:** Petek Şarlak Konya, Mehmet Nuri Konya, Bilge Kağan Yılmaz, Elif Kaga, Sadık Kaga, Yeliz Çetinkol

## Abstract

**Background:** Local antibiotic applications have been used in osteomyelitis as an adjunctive treatment method. Biodegradable materials are also used for the same purpose by adding antibiotics. The fact that it does not require additional surgery to be removed is an important advantage. The current study is therefore intended to develop a biodegradable drug-loaded polymeric scaffold with a good antibiotic release that can be an alternative to antibiotic-impregnated bone cement.

**Methods:** A tissue scaffold containing poly (2-hydroxyethyl methacrylate) (PHEMA) was prepared in our laboratory and loaded with Ertapenem and Daptomycin(E&D) antibiotics.

To evaluate drug release kinetics, the absorbance values of the scaffold loaded with E&D were measured with the spectrometer.

For microbiological tests, (E&D) impregnated cement and scaffold, as well as the control scaffold and cement samples were investigated for their antibacterial activities on Staphylococcus aureus and Klebsiella pneumoniae strains using the disc diffusion method.

**Results:** The Daptomycin zone diameter in S. aureus ATCC strain was 17 mm, 24 mm for scaffold, and 22 mm for cement. Scaffold was found to be more effective than cement against S. aureus strain. On K. pneumoniae ATCC strain, it was found that this strain was resistant to Ertapenem, but the zone diameter was 21 mm for scaffold and 20 mm for cement. Ertapenem-loaded scaffold was found to be more effective than cement.

When we evaluated the release profiles, for the Daptomycin-loaded cement group, 98%of Daptomycin was cumulatively released within 30 min, and for the Daptomycin-loaded scaffold group, 100%of Daptomycin was cumulatively released in 6 days. To compare Ertapenem- loaded cement and scaffold, 98% of Ertapenem was cumulatively released within 10 min in the cement group. For the scaffold group, 100% of Ertapenem was cumulatively released in 17 days. It was found that e scaffold released the antibiotic more slowly and for a longer duration.

## Introduction

Osteomyelitis is defined as a progressive disease characterized by microorganisms causing damage to bone tissue through infectious and inflammatory processes [1]. Only a part of the bone may be involved or the cortex, medullary canal, periosteum and surrounding soft tissues may all be affected. While osteomyelitis could have a mortal course in the past, with improved surgical options, compliance with sterility and antibiotic applications, it has been reduced to a severe disease and deaths have been almost completely prevented. However, despite the advances in today’s drug technologies and surgical treatments, it is difficult to reach the desired level of success [2].

The microorganisms causing osteomyelitis may vary depending on the age of the patient and the underlying diseases that s/he has. *S.aureus* stands out as the most frequently isolated pathogen in all types of osteomyelitis. *Coagulase negative staphylococci* are frequently isolated in infections due to foreign bodies such as implants and prostheses [3]. In nosocomial infections and in elderly and immunosuppressed patients, *P. aeruginosa*, *K. pneumonia* and other gram negative organisms are the responsible agents. Streptococci, anaerobic bacteria, *Pasteurella multicoda* and *Eikenella corrodens* are frequently isolated from human and animal bites. Diabetic foot and decubitus ulcer osteomyelitis is a polymicrobial infection [4]. In addition to gram negative agents, *Bartonella henselae, Aspergillus* species*, Mycobacterium avium* or *Candida albicans* have been reported in immunocompromised patients [5].

According to Cierny-Mader, two-stage treatment is applied in osteomyelitis surgery. The first stage involves radical debridement of dead bone and poorly vascularized soft tissues and systemic antibiotic therapy with placement of antibiotic-impregnated cement (AIC) (beads, rods, nails or blocks) in the resulting bone defect. In the second stage, after 6–8 weeks, the AIC is removed and the bone defect is filled with different methods [6]. AIC beads are used to sterilize and temporarily fill the dead cavity and are usually removed within 2–4 weeks [7]. The most commonly used antibiotics in these beads are cefazolin, moxalactam, cefotaxime, tobramycin, gentamicin, vancomycin and ticarcillin [8]. The disadvantage of this method is the need for repeated surgeries such as bone grafting or bone lengthening surgeries to remove the cement and fill the bone defects. Due to these shortcomings, bioeliminable carrier systems have started to be developed. These methods aim to achieve the high local antimicrobial concentrations required in the treatment of chronic osteomyelitis while avoiding the need for surgical removal of the implant. In addition, bioeliminable materials eliminate dead cavities in the bone and accelerate the healing of local bone tissue [9].

Antibiotics used systemically in the treatment of osteomyelitis have limited efficacy because they do not penetrate well into cavities filled with dead bone and necrotic content. Since these antibiotics are given in very high doses and for a long time for the same reasons, they have the disadvantages of side effects, systemic toxicity, long hospital stay, high cost, and the need for additional surgery. For this reason, research on local carrier systems and local antibiotic applications is increasing in number day by day [11].

In this study, we aimed to evaluate the antimicrobial efficacy of antibiotics implanted in a poly (2-hydroxyethyl methacrylate) (PHEMA)-based, bioeliminable, drug-loaded polymeric tissue scaffold with good antibiotic release, which can be an alternative to AIC.

Besides being a biocompatible and non-toxic polymer, PHEMA is a biomaterial used in many applications such as bone tissue regeneration, contact lenses, wound dressings. In such applications they are used in combination with crosslinkers such as ethylene glycol dimethylacrylate (EGDMA) for network formation. Such chemical cross-linked network structures of PHEMA are not bioabsorbable [12].

Bioabsorbability is a property that depends on the molecular weight and molecular structure of the polymer. Bioabsorbable polymers are first broken down into short chains or monomers in the organism and then these short chains and monomers are metabolized in the body or eliminated through the excretory system. Biocompatible polymers, on the other hand, when produced at the appropriate molecular weight (below the renal filtration threshold: 45 kDa) can be eliminated from the body [13]. Such structures are characterized as bioeliminable structures [14].

In this study, PHEMA-based and drug-loaded tissue scaffolds, which provide the advantage of bio-elimination and slow drug release, were developed as an alternative to routinely used cross-linked PMMA structures. For this purpose, tissue scaffolds were produced with very small molecular weight (average Mn: 530 g/mol) PHEMA chains by solvent casting without the use of crosslinkers [15].

## Materials and Methods

Financial support for the study was received from Afyonkarahisar University of Health Sciences Scientific Research Projects Unit with the project number 22.GENEL.016.

### Chemicals and equipment

Sodium metabisulfite was purchased from Acros organics, potassium persulfate from Akbel and 2-hydroxyethyl methacrylate (HEMA) from TCI companies. Diethyl ether and ethanol used in the polymerization stage and tissue scaffold production were supplied from the Merck company. Lyophilized Daptomycin was purchased from Novartis Sağlık, Gıda ve Tarım Ürünleri San. Tic. A.Ş. and lyophilized Ertapanem from Merck İlaç Ecza ve Kimya Ticaret AŞ. The surface morphology of the scaffolds was analyzed using LEO 1430 VP model scanning electron microscope (SEM) and BAL-TEC gold coating devices.

### PHEMA synthesis

For the synthesis of PHEMA the method in the literature was used [16]. First, 29 mg of potassium persulfate (0.1 mmol) and 29 mg of sodium metabisulfite (0.15 mmol) were dissolved homogeneously in 20 mL of 66.3% (v/v) ethanol/water mixture. Then 5 g of 2- hydroxyethyl methacrylate (HEMA) monomer (38.4 mmol) was added and the reaction flask was sealed with septum. O_2_ in the reaction medium was purged with high purity N_2_ gas for 15 min. The reaction was continued on a magnetic stirrer at room temperature for 6 h. The polymerization was then stopped by exposing the reaction to an open atmosphere. The polymer solution was dropped into 200 mL of cold ether stirred on a magnetic stirrer in an erlenmeyer to precipitate and purify PHEMA. The precipitated polymer was redissolved in 3 mL of ethanol and dropped into 200 mL of cold ether to dissolve again. This process was repeated once more and the polymer was purified by a total of three precipitations (Mn: 530 g/mol PDI: 1.2).

### Tissue scaffold production

Tissue scaffolds were produced using the solvent casting method as follows:

#### Ertapenem-loaded tissue scaffolds

200 mg PHEMA and 50 mg Ertapenem were dissolved in 700 µL ethanol/water mixture (6:1) by vortexing. In order to produce disk-shaped tissue scaffolds for cell tests and microbiological tests, 50 µL of this mixture was dropped onto a glass surface and dried in an oven at 25°C for 24 h.

#### Daptomycin-loaded tissue scaffolds

200 mg PHEMA and 50 mg Daptomycin were dissolved in 700 µL ethanol/water mixture (6:1) by vortexing. In order to produce disk-shaped tissue scaffolds for cell tests and microbiological tests, 50 µL of this mixture was dropped onto a glass surface and dried in an oven at 25°C for 24 h.

#### Drug-free tissue scaffolds

250 mg PHEMA was dissolved in 700 µL ethanol/water mixture (6:1) by vortexing. In order to produce disk-shaped tissue scaffolds for cell tests and microbiological tests, 50 µL of this mixture was dropped onto a glass surface and dried in an oven at 25°C for 24 h.

### SEM analysis of the tissue scaffolds

The surface morphology and pore geometries of drug-loaded and unloaded scaffolds were analyzed by SEM under vacuum.

After drying at 25°C for 24 h, the samples were coated using a BAL-TEC surface coating device to increase the surface conductivity. The coating process was carried out by sputtering. Gold plating was performed under 4x10^-2^ vacuum by applying 100 mA current for 20 s. The surface morphology of the scaffolds was then examined with a LEO 1430 VP model SEM device. Images were acquired in secondary electrons mode. Analysis was performed with an acceleration voltage of 20 kV.

### Cell culture: Proliferation test

Human osteoblast cell line (hFOB 1.19) was used in the study. Cells were incubated in 10% FBS, 100 U/mL penicillin-streptomycin supplemented media at 37°C and 5% CO_2_. First, the dose-dependent antiproliferative effects of the antibacterial drugs Ertapenem and Daptomycin were tested. Cells (5 × 10^3^ cells/well) were seeded in 96 well plates the night before. The next day, the medium of the cells was replaced with medium containing the drugs prepared at specific concentrations (0.01, 0.05, 0.5, 1.0, 3.0 mg/ml) and incubated for 48 h under cell culture conditions. After incubation, cells were incubated with 0.5 mg/ml MTT (dimethylthiazol diphenyltetrazolium bromide) reagent in serum-free medium for 4 h and then formazan crystals were dissolved with dimethyl sulfoxide. After treatment, absorbances were measured at 520 nm for each well using a plate reader (Thermosciantific, Multiscan FC). At the same time, the antiproliferative effects of scaffold, drug-loaded scaffold, cement and drug-loaded cement on osteoblast cells were investigated. Cells (5 × 10^4^ cells/well) were seeded in 24 well plates and allowed to adhere for 24 h. The next day, the scaffold, drug-loaded scaffold and cement and drug-loaded cement were placed on the attached cells in the wells and incubated under culture conditions for 48 h. After incubation, MTT assay was performed as described above.

### Release kinetics

Daptomycin and Ertapenem (20% (w/w)) were loaded onto polymeric tissue scaffolds to determine drug release kinetics. Each of the samples (10 mg) was then immersed in 10 ml of phosphate buffered saline and shaken in an incubator shaker at 37°C. At certain time points (0 h, 2 h, 24 h, 48 h, 72 h), absorbance values were measured by UV-VIS Spectrometer. Measurements were performed by spectrophotometry (Shimadzu, uv-1280) using wavelengths of 364 nm and 300 nm for Daptomycin and Ertapenem, respectively.

### Antimicrobial tests

Two different tissue scaffolds (PHEMA and PMMA) to which Ertapenem and Daptomycin were loaded together with their control samples were sent to Afyonkarahisar University of Health Sciences Health Application and Research Center Microbiology Laboratory under sterile conditions. The antibacterial activity of the scaffolds on *Staphylococcus aureus* ATCC 29213 and *Klebsiella pneumoniae* ATCC BAA-2814 strains was investigated by disk diffusion method.

### Preparation of the bacterial suspension at 0.5 McFarland (10^8^ cfu/ml) turbidity

A bacterial suspension was prepared in Mueller–Hinton broth (MHB) at 0.5 McFarland turbidity. For this purpose, 1–2 colonies of pure colony passaged bacteria were taken to prepare solutions at the 0.5 McFarland standard in a densitometer (BioMerieux, France).

### Obtaining 10^6^ cfu/ml concentrations of bacteria by serial dilutions

One ml of the original sample (10^8^ cfu/ml) was taken and added to 9 ml of MHB and mixed. The new sample had a cell concentration (cell count/ml) of 1/10 of the original sample. This process was repeated and serial dilutions were performed to obtain a bacterial concentration of 10^6^ cfu/ml. This step was performed separately for *Staphylococcus aureus* ATCC 29213 and *Klebsiella pneumoniae* ATCC BAA-2814 strains.

### Determination of antimicrobial activity of the scaffolds by disk diffusion method

Daptomycin and Ertapenem disks and the scaffold and cement controls were examined for their antimicrobial efficacy by disk diffusion method. 0.1 ml of the 10^6^ ml bacterial suspension was taken onto the blood agar and spread evenly over the entire surface of the medium with the help of a pipette. Daptomycin and Ertapenem were placed separately on each plate, and the Daptomycin- and Ertapenem-loaded scaffolds and cements were also placed onto plate using a sterile forceps. The media were incubated in an oven at 37°C for 24 h. One day later, the zone diameters on the plates removed from the oven were measured with a ruler and recorded.

## Results

### Production of tissue scaffolds

The dried tissue scaffolds prepared in the form of disks for cell culture and microbiological analyses are shown in Figure 1.

**Figure 1:**
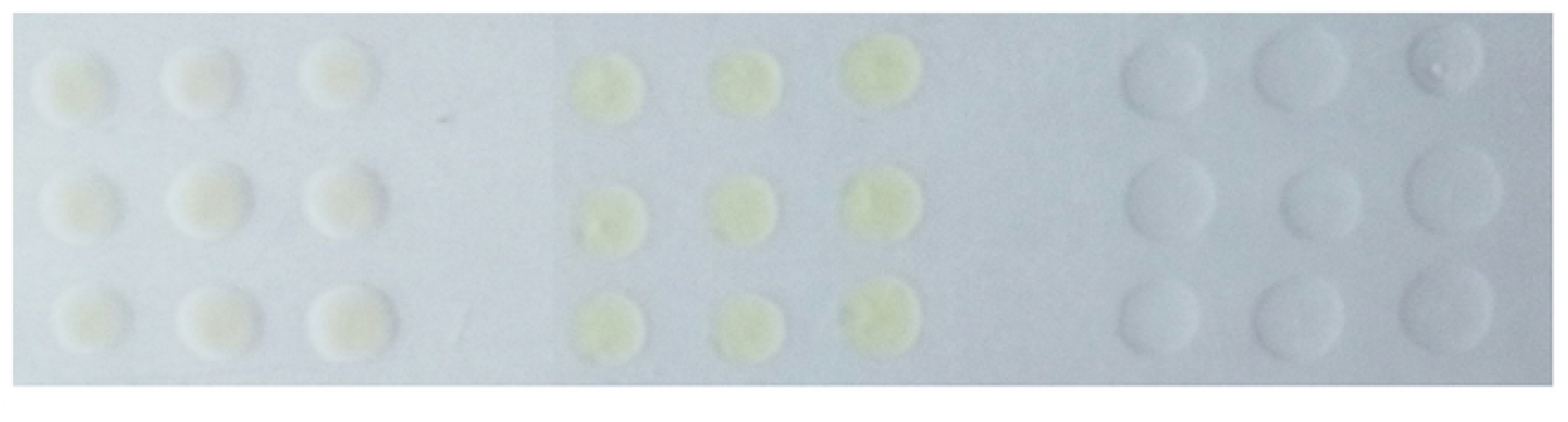
Ertapenem-loaded (left), Daptomycin-loaded (center) and drug-free (right) tissue scaffolds.

Drug-loaded tissue scaffolds were prepared with 20% drug load and were macroscopically equivalent in size and appearance.

### SEM analysis of the tissue scaffolds

SEM images of Ertapenem-loaded scaffold are shown in Figure 2a, Daptomycin-loaded scaffold in Figure 2b and drug-free scaffold in Figure 2c.

**Figure 2:**
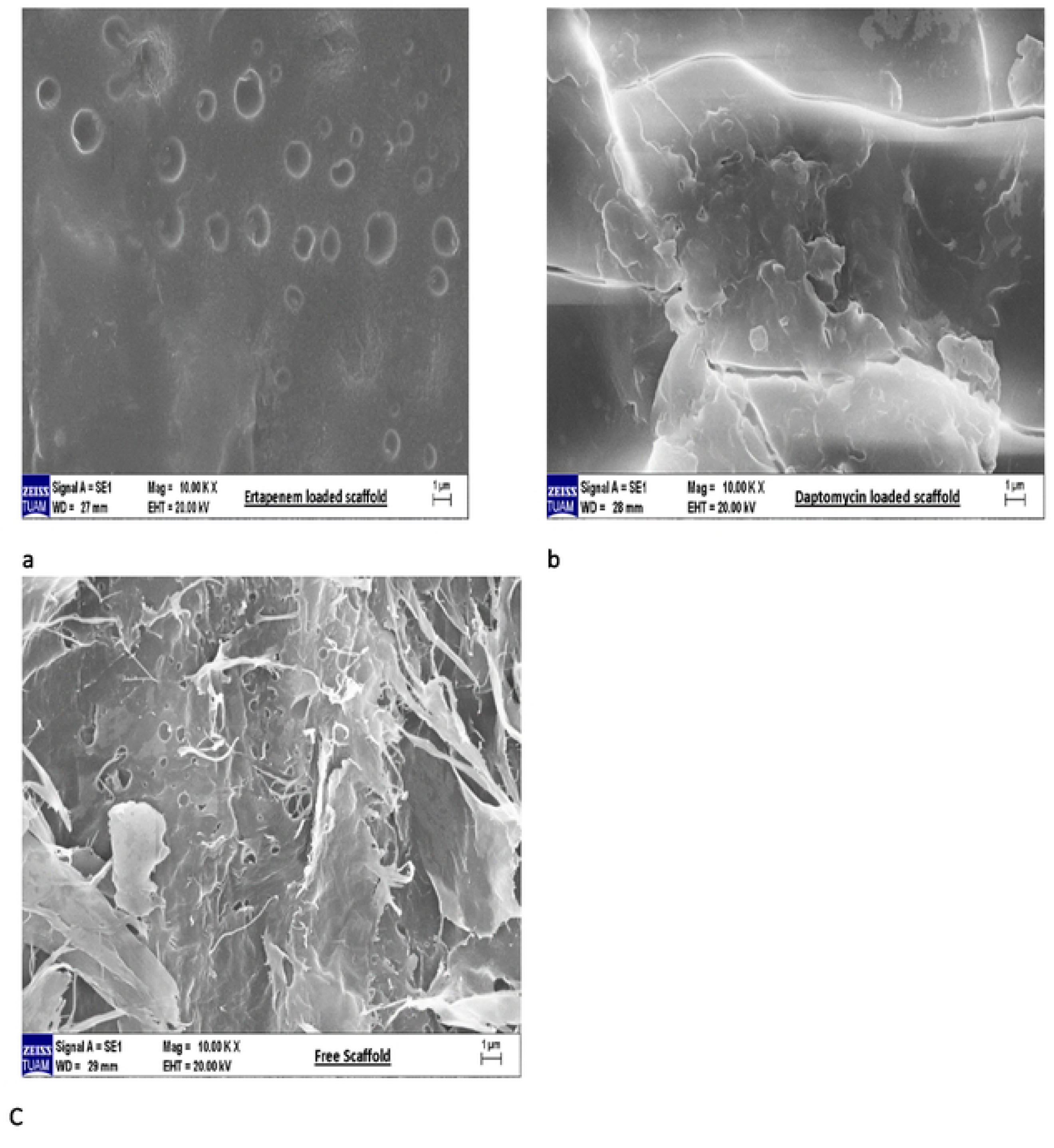
Ertapenem-loaded (left), Daptomycin-loaded (center) and drug-free (right) tissue scaffolds.

When the surface morphology of the scaffolds is examined, it can be said that each of them has a different surface morphology. The drug-loaded scaffolds show a more integrated structure than the drug-free scaffolds, whereas the drug-free scaffolds have a surface morphology with fibrillar extensions with more surface area.

### Cell proliferation assay

The antiproliferative effects of Ertapenem and Daptomycin on osteoblast cells were examined. The drugs showed a dose-dependent antiproliferative effect. When both drugs were applied at doses of 0.01 mg/ml, 0.05 mg/ml and 0.5 mg/ml, cell viability rates were 90%, whereas the cells where the drugs were applied at increasing doses (1 mg/ml and 3 mg/ml) had a rate of cell death of approximately 40% (Figure 3).

**Figure 3:**
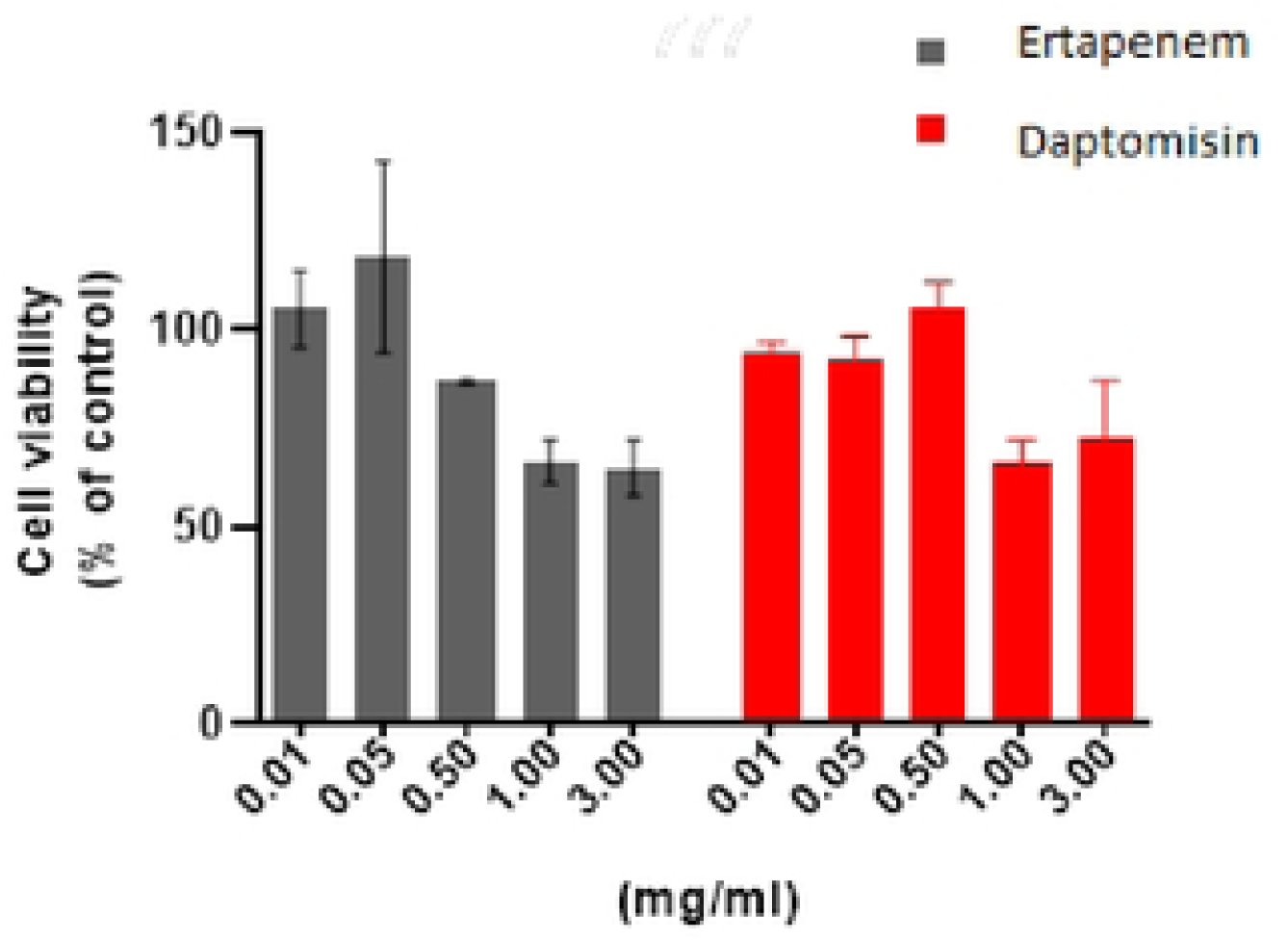
Scanning electron microscope (SEM) images a) SEM image of the Ertapenem-loaded scaffold b) SEM image of the Daptomycin-loaded scaffold c) SEM image of the drug-free scaffold.

The antiproliferative effect of Ertapenem- and Daptomycin-loaded scaffold and cement on osteoblast cells was investigated. Approximately 90% of cell viability was detected in the Ertapenem-loaded scaffold and Ertapenem-loaded cement group. In the Daptomycin-loaded scaffold and Daptomycin-loaded cement group, cell viability was approximately 50% (Figure 4).

**Figure 4:**
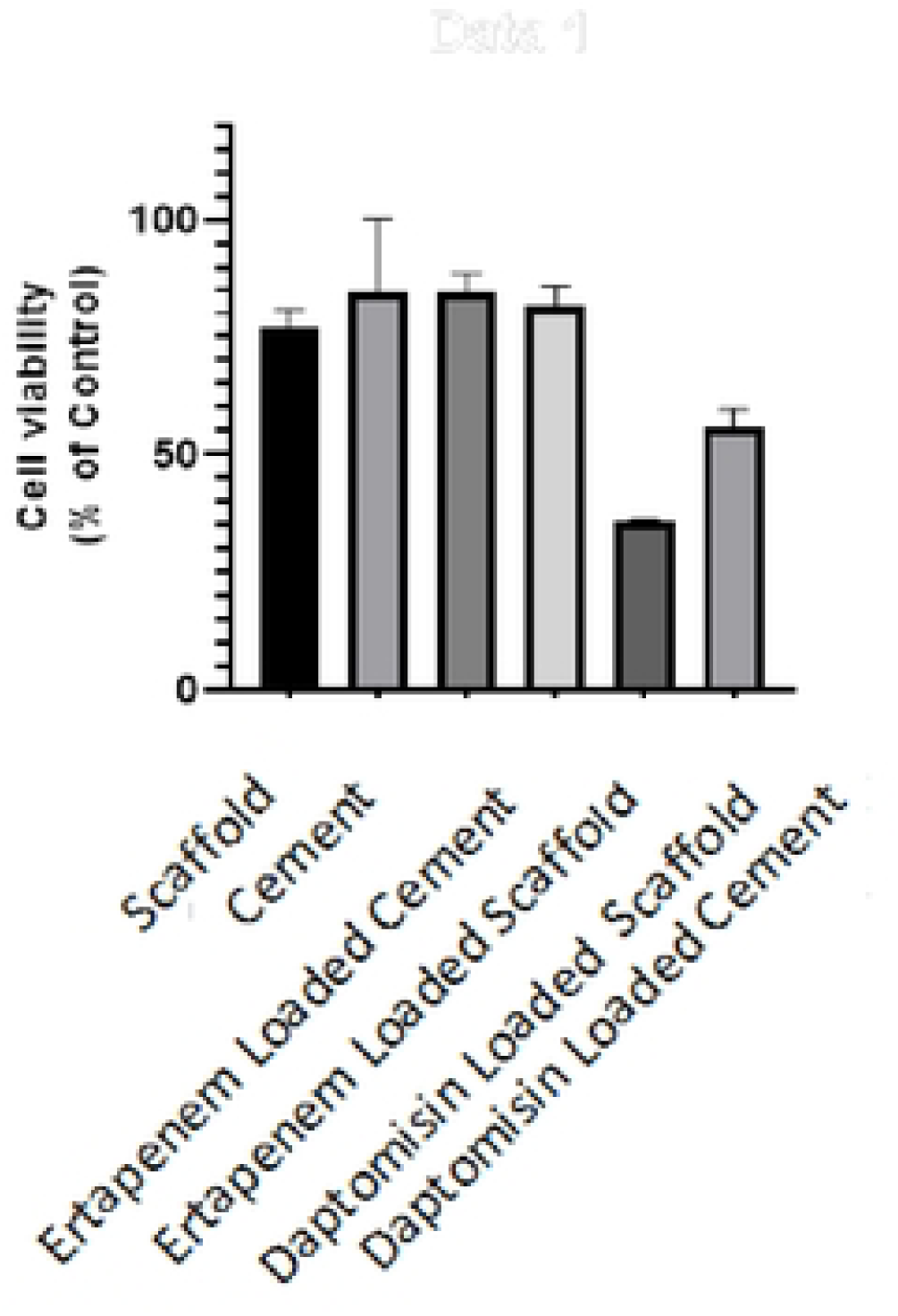
Effects of Ertapenem and Daptomycin on cell viability measured by dimethylthiazol diphenyltetrazolium bromide assay: Osteoblast cells were treated with Ertapenem and Daptomycin at various concentrations (0.01–3 mg/ml) for 48 h. Results are represented as the mean ± standard deviation of triplicate independent experiment.

Daptomycin and Ertapenem were loaded into both scaffold and cement to demonstrate the comparative release profile of scaffold and cement. In the Daptomycin-loaded cement group, approximately 98% of Daptomycin was cumulatively released within 30 min. In the Daptomycin-loaded scaffold group, approximately 100% of Daptomycin was cumulatively released in 6 days (Figure 5a). When Ertapenem-loaded cement and Ertapenem-loaded scaffold were compared, approximately 98% of Ertapenem was cumulatively released within 10 min in the Ertapenem-loaded cement group. In the Ertapenem-loaded scaffold group, approximately 100% of Ertapenem was cumulatively released in 17 days (Figure 5b).

**Figure 5:**
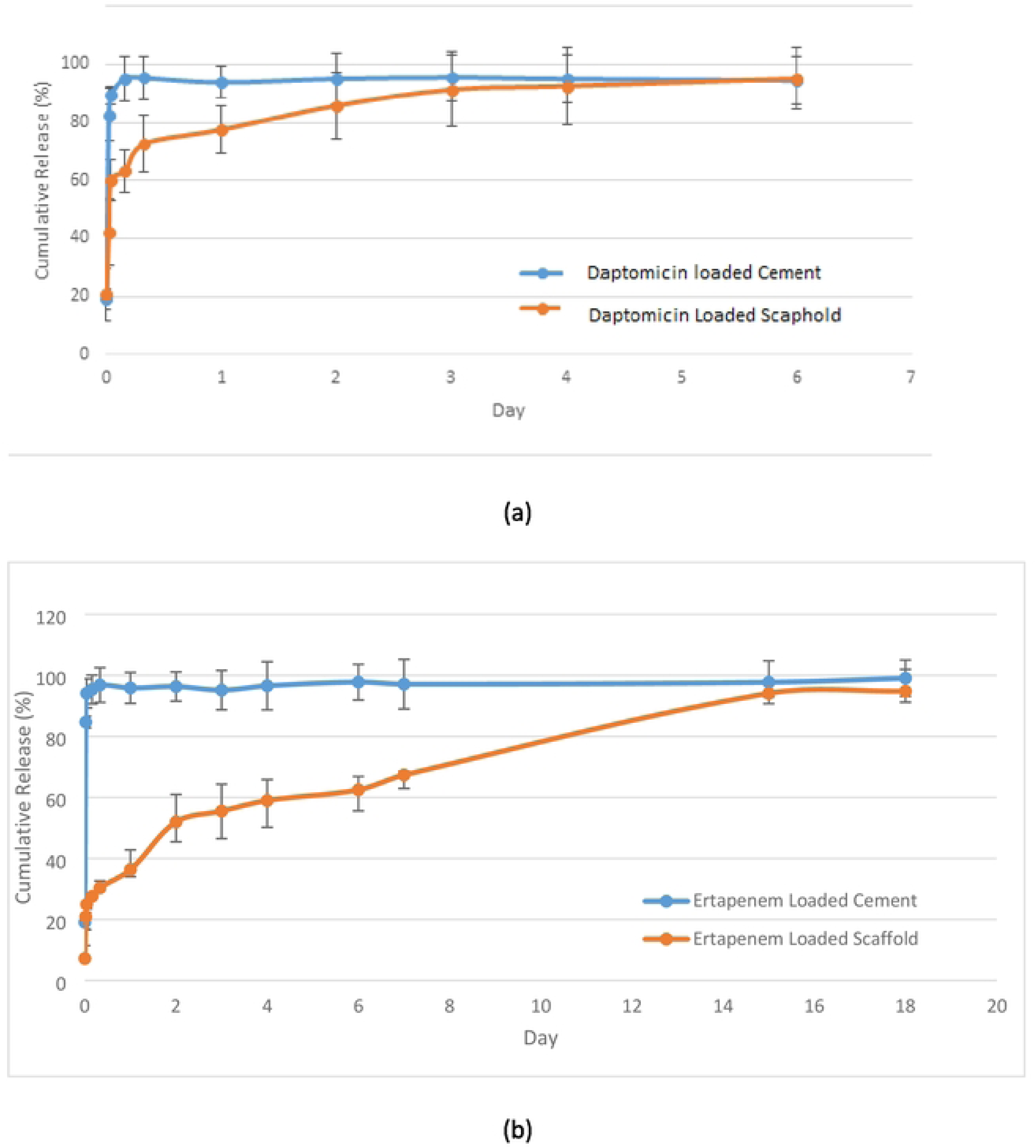
Dimethylthiazol diphenyltetrazolium bromide assay for osteoblast cell proliferation on polymeric scaffold, cement, Ertapenem-loaded scaffold, Ertapenem-loaded cement, Daptomycin-loaded scaffold, Daptomycin-loaded cement. Bar expresses mean ± standard deviation of triplicate independent experiments.

**Figure 6:** Release profile of Daptomycin (a) and Ertapenem (b) on polymeric scaffold. 20% (w/w) Daptomycin and Entarpenem were loaded in each scaffold, and the amount of released drugs were quantified by spectrophotometry. Error bars indicate standard deviation, n = 3.

### Microbiological results

The efficacy of antibiotic-loaded scaffold and cement on the two most common microorganisms was investigated by disk diffusion method (Table 1). In *S. aureus* ATCC 29213 strain, Daptomycin zone diameter was 17 mm, whereas this value was 24 mm for scaffold and 22 mm for cement. Scaffold was found to be more effective than cement against *S. aureus* ATCC 29213 strain, whereas both building materials were more effective than Daptomycin disk.

**Table 1:** Comparison of the efficacies of scaffold and cement with those of antibiotic discs according to the disk diffusion method performed.

When *K. pneumoniae* ATCC strain was analyzed, it was found that this strain was resistant to Ertapenem, but the zone diameter was 21 mm for the scaffold and 20 mm for the cement. Of the Ertapenem-impregnated building materials, scaffold was found to be more effective than bone cement, and both scaffold and bone cement were also found to be more effective than the Ertapenem disk.

As a result, the antimicrobial efficacy of scaffold was found to be better than cement according to the disk diffusion method.

## Discussion

Today, polymethyl methacrylate (PMMA) cement is considered the gold standard antibiotic carrier in the treatment of osteomyelitis [17]. The main advantage of local antibiotic application with cementation is the high local antibiotic concentrations achieved, minimizing the risk of systemic toxicity, preventing emerging antibiotic resistance. There are many studies in the literature with PMMA cement. Keeling et al. experimentally demonstrated that the bacterial biofilm layer can be reduced with vancomycin and Daptomycin impregnated cement in their animal study [18]. In a study by Bor et al. on 16 patients with chronic osteomyelitis, radical debridement was performed and AIC was applied to the bone defect. No recurrence was detected during a mean follow-up period of 6 years. However, in this study, the removal of cement with an additional operation is considered as a disadvantage [19]. Therefore, in recent years, the development of bioeliminable tissue scaffolds that can be used instead of cement has gained momentum. With bioeliminable scaffolds, it is aimed to utilize both the local release effect of antibiotic-loaded scaffolds and to avoid the need for a second surgery. Sambri et al. developed a bioeliminable scaffold consisting of a combination of calcium phosphate and nanocrystalline hydroxyapatite (HA) and investigated its local effect by impregnating the scaffold with antibiotics in animal models of osteomyelitis. In this study with single-stage debridement, favorable results were obtained despite the small number of subjects and short follow-up period [20]. Alegrete et al. used a ceramic biomaterial loaded with antibiotics in the treatment of osteomyelitis in animal models. The ceramic biomaterial used was found to be effective in the treatment of osteomyelitis while supporting bone formation and osteointegration [21].

In this study, we have produced a bioeliminable tissue scaffold containing PHEMA as an alternative to AIC. We loaded Ertapenem and Daptomycin on the scaffold we produced and evaluated the release kinetics and antimicrobial activity in an in vitro environment.

The selection of the right antibiotic-loaded onto the scaffolds, the way it is combined with cement, the amount of antibiotic, the release time and form of the drug into the surrounding tissues are of great importance in its use. The selected antibiotic should contain a broad antibacterial spectrum, be water-soluble, nonallergenic, and thermostable [22]. The most commonly used antibiotics in studies are cefazolin, moxalactam, cefotaxime, tobramycin, gentamycin, vancomycin and ticarcillin [8]. Therate of methicillin resistance in *S. aureus*, the most common causative agent of osteomyelitis, is increasing day by day. In a study by Ahmed et al. in patients with soft tissue infections, *MRSA (Methylicin-resistant S.aureus)* has been detected in 89.9% of the *S. aureus* isolates [23]. Hence, the treatment options *MRSA* effective antibiotics should be considered. In our study, we added Daptomycin, which may be effective against possible gram positive agents, into the scaffold we produced. Daptomycin is an antibiotic with bactericidal activity against gram positive agents with MRSA activity [24]. *Klebsiella pneumoniae*is one of the most frequently isolated gram negative agents in cases of osteomyelitis. In studies conducted in osteomyelitis, cephalosporin resistance is around 80% in *K. pneumonia* strains [25]. Carbapenems remain a good treatment option for the Enterobacteriaceae family that produces extended-spectrum beta-lactamase. In our study, we added Ertapenem, an effective carbapenem against possible gram negative agents, into the scaffold we produced.

In the literature, studies have been conducted on the antimicrobial activity of many tissue scaffolds on these microorganisms. Campos et al. compared vancomycin-loaded cement and calcium sulfate-containing tissue scaffolds on *S.aureus* strains. This study showed that both calcium sulfate-containing tissue scaffolds and cement release antibiotics for up to 2 weeks and act as good antibiotic carriers. However, scaffolds containing calcium sulfate showed greater antimicrobial release potential than cement. Therefore, it was concluded that tissue scaffolds containing antimicrobial-impregnated calcium sulfate showed high bioactivity to kill biofilm cells. In this study, the efficacy on gram negative resistant agents was not evaluated, which is a limitation of the study [26].

In our study, the efficacy of antibiotic-impregnated scaffold and cement on both gram negative and gram positive microorganisms was investigated using the disk diffusion method.

Daptomycin zone diameter in *S. aureus* ATCC strain was 17 mm, whereas this value was 24 mm for scaffold and 22 mm for cement. Scaffold was found to be more effective than cement against *S. aureus* ATCC strain, whereas both building materials were more effective than Daptomycin disk.

On *K. pneumoniae* ATCC strain, it was found that this strain was resistant to Ertapenem, but the zone diameter was 21 mm for scaffold and 20 mm for cement. As a result, Ertapenem- loaded scaffold was more effective than cement and both cement and Ertapenem-loaded scaffold were more effective than Ertapenem disk. Accordingly, we found that the antimicrobial efficacy of the scaffold we developed was better than cement. Therefore, we think that scaffold is a good alternative to cement.

We also evaluated the controlled release profile of the scaffold loaded with Daptomycin and Ertapenem in comparison with the release profile of cementum, the most commonly used antibiotic carrier in daily practice. In the Daptomycin-loaded cementum group, approximately 98% of Daptomycin was cumulatively released within 30 min. In the Daptomycin-loaded scaffold group, approximately 100% of Daptomycin was cumulatively released in 6 days. To compare Ertapenem-loaded cement and scaffold, approximately 98% of Ertapenem was cumulatively released within 10 min in the Ertapenem-loaded cement group. For the Ertapenem-loaded scaffold group, approximately 100% of Ertapenem was cumulatively released in 17 days. As a result, we found that the scaffold we developed released the antibiotic more slowly and during a longer duration. It is thought that a bioeliminable scaffold that offers controlled and slow drug release may be more effective on biofilm and may be more successful in the treatment of osteomyelitis.

## Conclusion

In our study, the scaffold we produced was compared with cement, which we frequently use in daily practice, and it was concluded that the scaffold has better drug release and antimicrobial efficacy. In addition, our scaffold is more advantageous than cement because it is bioeliminable. Thus, a second surgical intervention will not be necessary and possible mortality and morbidities will be prevented. Because of all these features, scaffold seems promising in the local treatment of osteomyelitis. Our study was conducted under *in vitro* conditions and further extensive studies with animal experiments are needed.

## Data Availability

Data cannot be shared publicly because of [Afyonkarahisar Health Science University.

## Supplementary Materials

## Author Contributions

Conceptualization, PŞK and MNK; Methodology, EK; Software, SK; Validation EK,SK Formal Analysis, SK; Investigation,BKY ; Resources, PK, BKY; Data Curation, YC.; Writing – Original Draft Preparation, PŞK; Writing – Review & Editing, YC, MNK; Visualization, SK; Supervision, YC; Project Administration, PŞK; Funding Acquisition, PŞK”,

## Institutional Review Board Statement

Afyonkarahisar Health Science University, Institutional Review Board,meeting date: 13.05.2022, No: 2022/6.

## Informed Consent Statement

Consent is not required as it is an experimental study.

## Data Availability Statement

The study did not report any data.

## Acknowledgments

Author thanks Enago Corp. for English Translation and Formatting.

## References

1. Demir, B.; Gursu, S.; Oke, R.; Konya, N.M.; Ozturk, K.; Sahin, V. Shortening and secondary relengthening for chronically infected tibial pseudarthroses with poor soft tissues. J. Orthop. Sci. 2009, 14, 525–534. DOI:10.1007/s00776-009-1364-5.

2. Schmitt SK. Osteomyelitis. Infect Dis Clin North Am. 2017 Jun;31(2):325–338. doi: 10.1016/j.idc.2017.01.010. PMID: 28483044.

3. 3. García Del Pozo E, Collazos J, Cartón JA, Camporro D, Asensi V. Bacterial osteomyelitis: microbiological, clinical, therapeutic, and evolutive characteristics of 344 episodes. Rev Esp Quimioter. 2018 Jun;31(3):217–225. Epub 2018 May 11. PMID: 29756429; PMCID: PMC6166254.

4. Oliver TI, Mutluoglu M. Diabetic Foot Ulcer. 2022 Aug 8. In: StatPearls [Internet]. Treasure Island (FL): StatPearls Publishing; 2023 Jan–.

5. Bariteau JT, Waryasz GR, McDonnell M, Fischer SA, Hayda RA, Born CT. Fungal osteomyelitis and septic arthritis. J Am Acad Orthop Surg. 2014 Jun;22(6):390–401. doi: 10.5435/JAAOS-22-06-390.

6. Subramanyam KN, Mundargi AV, Prabhu MV, Gopakumar KU, Gowda DSA, Reddy DR. Surgical management of chronic osteomyelitis: Organisms, recurrence and treatment outcome. Chin J Traumatol. 2023 Jul;26(4):228–235. doi: 10.1016/j.cjtee.2023.01.003.

7. Flores MJ, Brown KE, Morshed S, Shearer DW. Evidence for Local Antibiotics in the Prevention of Infection in Orthopaedic Trauma. J Clin Med. 2022 Dec 16;11(24):7461. doi: 10.3390/jcm11247461.

8. Jiamton C, Apivatgaroon A, Aunaramwat S, et al Efficacy and Safety of Antibiotic Impregnated Microporous Nanohydroxyapatite Beads for Chronic Osteomyelitis Treatment: A Multicenter, Open-Label, Prospective Cohort Study. Antibiotics (Basel). 2023 Jun 15;12(6):1049. doi: 10.3390/antibiotics12061049.

9. Guo X, Song P, Li F, Yan Q, Bai Y, He J, Che Q, Cao H, Guo J, Su Z. Research Progress of Design Drugs and Composite Biomaterials in Bone Tissue Engineering. Int J Nanomedicine. 2023 Jul 1;18:3595–3622. doi: 10.2147/IJN.S415666.

10. Morinaga S, Yamamoto N, Tokoro M, Hayashi K, Takeuchi A, Miwa S, Igarashi K, Taniguchi Y, Asano Y, Nojima T, Tsuchiya H. Antibacterial effect and biological reaction of calcium phosphate cement impregnated with iodine for use in bone defects. J Biomater Appl. 2023 May;37(10):1716–1723. doi: 10.1177/08853282231164827.

11. Ferguson J, Bourget-Murray J, Stubbs D, McNally M, Hotchen AJ. A comparison of clinical and radiological outcomes between two different biodegradable local antibiotic carriers used in the single-stage surgical management of long bone osteomyelitis. Bone Joint Res. 2023 Jul 4;12(7):412–422. doi: 10.1302/2046-3758.127.BJR-2022-0305.R2.

12. Zare M, Bigham A, Zare M, Luo H, Rezvani Ghomi E, Ramakrishna S. pHEMA: An Overview for Biomedical Applications. Int J Mol Sci. 2021 Jun 15;22(12):6376. doi: 10.3390/ijms22126376.

13. Chen K, Cai H, Zhang H, Zhu H, Gu Z, Gong Q, Luo K. Stimuli-responsive poly-mer- doxorubicin conjugate: Antitumor mechanism and potential as nano-prodrug. Acta Biomater. 2019 Jan 15;84:339–355. doi: 10.1016/j.actbio.2018.11.050.

14. Gao D, Xu H, Philbert MA, Kopelman R. Bioeliminable nanohydrogels for drug delivery. Nano Lett. 2008 Oct;8(10):3320–4. doi: 10.1021/nl8017274.

15. Borbolla-Jiménez FV, Peña-Corona SI et al. Films for Wound Healing Fabricated Us-ing a Solvent Casting Technique. Pharmaceutics. 2023 Jul 9;15(7):1914. doi: 10.3390/pharmaceutics15071914.

16. Zhang B, Lalani R, Cheng F, Liu Q, Liu L. Dual-functional electrospun poly(2- hydroxyethyl methacrylate). J Biomed Mater Res A. 2011 Dec 1;99(3):455–66. doi: 10.1002/jbm.a.33205.

17. Al Thaher Y, Khalil R, Abdelghany S, Salem MS. Antimicrobial PMMA Bone Cement Containing Long Releasing Multi-Walled Carbon Nanotubes. Nanomaterials (Basel). 2022 Apr 18;12(8):1381. doi: 10.3390/nano12081381. PMID: 35458089; PMCID: PMC9026701.

18. Keeling WB, Myers AR, Stone PA, Heller L, Widen R, Back MR, Johnson BL, Bandyk DF, Shames ML. Regional antibiotic delivery for the treatment of experimental prosthetic graft infections. J Surg Res. 2009 Dec;157(2):223–6. doi: 10.1016/j.jss.2008.06.050.

19. Bor N, Dujovny E, Rinat B, Rozen N, Rubin G. Treatment of chronic osteomyelitis with antibiotic-impregnated polymethyl methacrylate (PMMA) - the Cierny approach: is the second stage necessary? BMC Musculoskelet Disord. 2022 Jan 6;23(1):38. doi: 10.1186/s12891-021-04979-y.

20. Sambri A, Cevolani L, Passarino V, Bortoli M, Parisi SC, Fiore M, Campanacci L, Staals E, Donati DM, De Paolis M. Mid-Term Results of Single-Stage Surgery for Patients with Chronic Osteomyelitis Using Antibiotic-Loaded Resorbable PerOssal® Beads. Microorganisms. 2023 Jun 21;11(7):1623. doi: 10.3390/microorganisms11071623.

21. Alegrete, N.; Sousa, S.R.; Peleteiro, B.; Monteiro, F.J.; Gutierres, M. Local antibiotic delivery ceramic bone substitutes for the treatment of infected bone cavities and bone regen-eration: A systematic review on what we have learned from animal models. Materials (Basel). 2023, 16. DOI:10.3390/ma16062387.

22. Bistolfi A, Ferracini R, Albanese C, Vernè E, Miola M. PMMA-Based Bone Cements and the Problem of Joint Arthroplasty Infections: Status and New Perspectives. Materials (Basel). 2019 Dec 2;12(23):4002. doi: 10.3390/ma12234002.

23. Ahmed EF, Rasmi AH, Darwish AMA, Gad GFM. Prevalence and resistance profile of bacteria isolated from wound infections among a group of patients in upper Egypt: a descriptive cross-sectional study. BMC Res Notes. 2023 Jun 19;16(1):106. doi: 10.1186/s13104-023-06379-y.

24. García P, Moscoso M, Fernández MC, Fuentes-Valverde V, Pérez A, Bou G. Compari-son of the in vivo efficacy of ceftaroline fosamil, vancomycin and daptomycin in a murine model of methicillin-resistant Staphylococcus aureus bacteraemia. Int J Antimicrob Agents. 2023 Jul;62(1):106836. doi: 10.1016/j.ijantimicag.2023.106836.

25. Shipitsyna IV, Osipova EV. Efficacy of cephalosporins against enterobacteria isolated from patients with chronic osteomyelitis. Klin Lab Diagn. 2022 Mar 25;67(3):158–162. Doi: 10.51620/0869-2084-2022-67-3-158-162

26. Batista Campos L, Kurihara MNL, Santos INM, Dos Reis FB, Salles MJ. In vitro elu-tion characteristics of antibiotic-loaded polymethylmethacrylate cement and a calcium sulfate bone substitute using staphylococci isolated from orthopedic implant-associated infections. J Biomed Mater Res B Appl Biomater. 2023 Jun;111(6):1318–1327. doi: 10.1002/jbm.b.35235.

